# Graphene nanoplatelet and Graphene oxide functionalization of face mask materials inhibits infectivity of trapped SARS-CoV-2

**DOI:** 10.1101/2020.09.16.20194316

**Authors:** Flavio De Maio, Valentina Palmieri, Gabriele Babini, Alberto Augello, Ivana Palucci, Giordano Perini, Alessandro Salustri, Marco De Spirito, Maurizio Sanguinetti, Giovanni Delogu, Laura Giorgia Rizzi, Giulio Cesareo, Patrick Soon-Shiong, Michela Sali, Massimiliano Papi

**Affiliations:** Dipartimento di Scienze di Laboratorio e Infettivologiche, Fondazione Policlinico Universitario “A. Gemelli” IRCSS, Rome, Italy; Dipartimento di Scienze biotecnologiche di base, cliniche intensivologiche e perioperatorie – Sezione di Microbiologia, Università Cattolica del Sacro Cuore, Rome, Italy; Dipartimento di Neuroscienze, Università Cattolica del Sacro Cuore, Rome, Italy; Fondazione Policlinico Universitario “A. Gemelli” IRCSS, Rome, Italy; Istituto dei Sistemi Complessi, CNR, Via dei Taurini 19, 00185 Rome, Italy; Dipartimento Scienze della Salute della Donna, del Bambino e di Sanità Pubblica, Fondazione Policlinico Universitario “A. Gemelli”, IRCCS, Rome, Italy; Mater Olbia Hospital, Olbia, Italy; Directa Plus S.p.A. c/o ComoNExT – Science and Technology Park, Lomazzo, Italy; Nantworks LLC, Culver City, California, USA

## Abstract

Recent advancements in bidimensional nanoparticles such as Graphene nanoplatelets (G) and the derivative Graphene oxide (GO) have the potential to meet the need for highly functional personal protective equipment (PPE) that confers increased protection against SARS-CoV-2 infection and the spread COVID-19. The ability of G and GO to interact with and bind microorganisms as well as RNA and DNA provides an opportunity to develop engineered textiles for use in PPE. The face masks widely used in health care and other high-risk settings for COVID transmission provide only a physical barrier that decreases likelihood of infection and do not inactivate the virus. Here, we show pre-incubation of viral particles with free GO inhibits SARS-CoV-2 infection of VERO cells. Highly relevant to PPE materials, when either polyurethane or cotton material was loaded with G or GO and culture medium containing SARS-CoV-2 viral particles either filtered through or incubated with the functionalized materials, the infectivity of the medium was nearly completely inhibited. The findings presented here constitute an important nanomaterials-based strategy to significantly increase face mask and other PPE efficacy in protection against the SARS-CoV-2 virus and COVID-19 that may be applicable to additional anti-SARS-CoV-2 measures including water filtration, air purification, and diagnostics.

**One Sentence Summary:** Cotton and polyurethane materials functionalized with bidimensional Graphene nanoplatelets trap SARS-CoV-2 and have the potential to reduce spread of COVID-19.

---

The emergence of Severe Acute Respiratory Syndrome Coronavirus 2 (SARS-CoV-2) and the resultant coronavirus infectious disease 2019 (COVID-19) pandemic has prompted the ubiquitous use of face masks (*1–4*) to curb transmission in clinical, public, and working contexts. A global shortage of medical-grade masks such as N95 masks has resulted in widespread use of cloth masks, including woven cotton masks, to reduce airborne transmission (*4*). While these masks offer some protection against viral transmission, given what will likely be continuing high demand for personal protective equipment (PPE) particularly due to the inability to avoid or difficulty in isolation of infectious persons that are contagious during the initial days of infection when symptoms are mildest or not present (*5, 6*), masks and PPE with better protective characteristics are needed.

To improve the protective properties of masks, nanomaterials scientists have proposed integration of virus-inactivating properties with standard propylene- and cloth-filtering properties with the goal of developing improved PPE (*7–10*).

In recent years, the bidimensional material Graphene nanoplatelet (G) and its derivatives have captured much attention due to their interactions with microorganisms (*11–13*). Pristine G is a single-atom-thick sheet of hexagonally arranged carbon atoms (*14*), while Graphene oxide (GO) is its oxidized form. Being a single layer of carbon atoms, G has an exceptionally high surface area and interacts uniquely with organisms with sizes in the order of hundreds of nanometers, i.e. bacteria and viruses (*10*). GO oxygen groups make its surface more hydrophilic compared to G and facilitate GO interaction with organic molecules when in solution (*15, 16*).

It has been demonstrated that bacteria that come into contact with the G surface lose integrity (*16, 17*) and that G has good viral inhibition capacity. Graphene interacts directly with viruses mainly by hydrogen bonding, electrostatic interactions, and redox reactions (*18*); and many G-derived materials have an intrinsic ability to adsorb charged lipids and destroy membranes (*19–21*) such as those of enveloped viruses like SARS-CoV-2. Furthermore, G itself can be additionally functionalized with antiviral particles and drugs (*22–29*).

In the studies described here, we first investigated the ability of GO in solution to bind and entrap SARS-CoV-2 viral particles by incubating ~10^5^ viral particles/mL with increasing concentrations of GO for 2 hours. Following incubation, solutions were centrifuged and supernatants used to incubate VERO (ATCC CCL-81) cells to measure the infectivity of the virus (Fig. 1A). As shown in Fig. 1B, incubation of SARS-CoV-2 viral particles in suspension with GO dramatically reduced viral infectivity even at the lowest concentration of GO (0.06 mg/mL), as reflected by decrease of the cytopathic effect of the virus and increased cell survival. GO also significantly reduced the load of viral particles as assessed by immunofluorescent labeling with an anti-SARS-CoV-2 spike protein antibody (Fig. 1C and 1E) and reduced cellular cytotoxicity as measured by both Crystal violet staining (Fig. 1D and 1F) and lactate dehydrogenase (LDH) release in the supernatant (Fig. 1G). These data demonstrate that water-soluble GO interacts with SARS-CoV-2 viral particles and reduces viral infectivity in the *in vitro* live virus model of SARS-CoV-2 infection of VERO cells.

**Figure 1.**
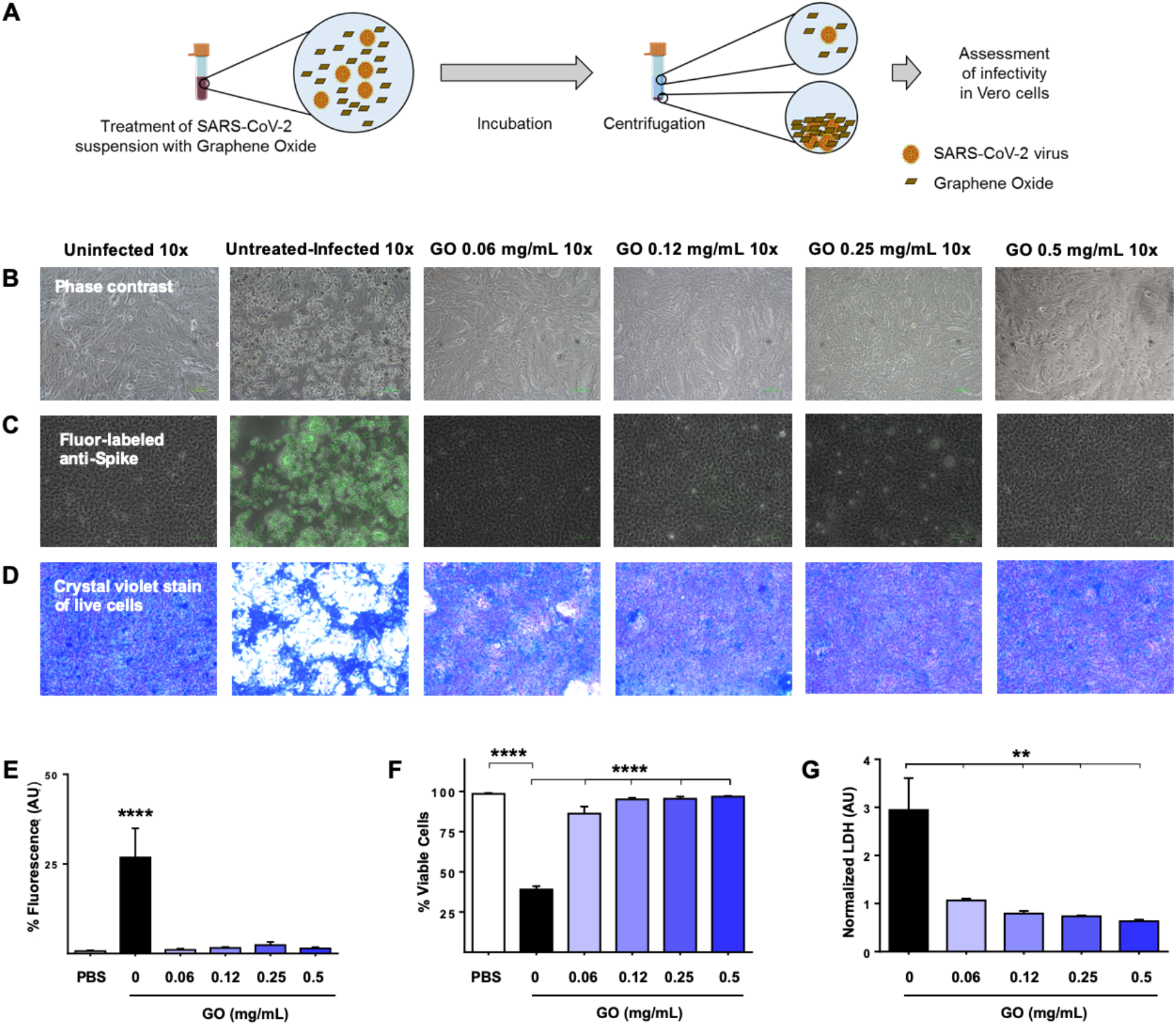
Graphene oxide (GO) entraps the SARS-CoV-2 virus and prevents infection. (A) A schematic representation of the experimental design to assess the ability of GO to trap virus in solution is shown. A SARS-CoV-2 clinical isolate was suspended in phosphate buffered saline (PBS) at ~10^5^ virus particles/mL and incubated with increasing concentrations of GO (0.06, 0.12, 0.25, and 0.5 mg/mL) or without GO (untreated) as positive control. Two hours later, GO was removed by centrifugation and supernatants used to infect VERO cells. Cell viability was monitored daily and representative images taken at 72 h post-infection by (B) light microscopy to visualize cell density or (C) fluorescent microscopy following immunofluorescent labeling of cells with an anti-viral spike (S) protein antibody. (D) Cell viability was also assayed with Crystal violet staining. (E) Fluorescence and (F) Crystal violet staining images were analyzed using ImageJ software to quantify infected cells and cytotoxicity, respectively. (G) Lactate dehydrogenase (LDH) was quantified in the cell supernatants to measure SARS-CoV-2-mediated cytotoxicity. Data were graphed as the mean with SD. All experiments were analyzed by using one-way ANOVA tests followed by Tukey’s correction (p < 0.05 = *; p < 0.01 = **; p < 0.001 = ***).

The ability of GO to trap pathogens was one of the first properties attributed to this nanomaterial and was reported to result in inhibition of macrophage infection by *Mycobacterium tuberculosis* (*30–32*), as well as bacteriostatic effects on both Gram-positive and Gram-negative bacteria (*29*).

In addition to the potential for integration of GO in PPE, the ability of GO to trap infectious viral particles provides an opportunity for the treatment of water effluents from hospitals and municipalities. This may be a critical application of GO technology, given that coronaviruses can maintain viability in sewage and hospital wastewater, and can persist in aquatic environments and wastewater treatment plants (*33*). With further development, GO technology could be applied to water treatment, air purification and – in combination with monitoring programs – has the potential to reduce environmental viral loads and secondary transmission.

In addition, the GO trapping effect can be used to concentrate analytes in solution, such as DNA, RNA, and viruses (*34*), a characteristic that can be leveraged in design of new diagnostic methods.

Here, in our next line of investigation, we sought to elucidate the potential to exploit the interaction of Graphene with viruses in the fabrication of masks by testing the efficacy of G and GO functionalization of cotton and non-woven, polyurethane (PU) material.

Design of an effective surgical face mask consists of the use of materials with different roles and properties, layered in a sequence to optimize protection. The first layer, in contact with the skin of the wearer, allows droplets to pass through and be absorbed in the adjacent hydrophilic layer, thereby keeping the skin dry (*35*). This second hydrophilic layer also absorbs and holds microorganisms in the mask, limiting their ability to spread to other people.

In contrast, the outside layer of the mask is typically a hydrophobic non-woven tissue sheet. Its low wettability prevents escape of fluids from the middle layer to the outside and at the same time stops entry of droplets from the exterior.

To assess if functionalization of cotton material or PU is feasible and can result in better protection against viral penetration, we first integrated G and GO into these materials. The functionalized materials were then characterized by scanning electron microscopy (SEM). As seen in Fig. 2, SEM imaging reveals platelets of G or GO distributed on fibers that are particularly apparent on PU.

**Figure 2.**
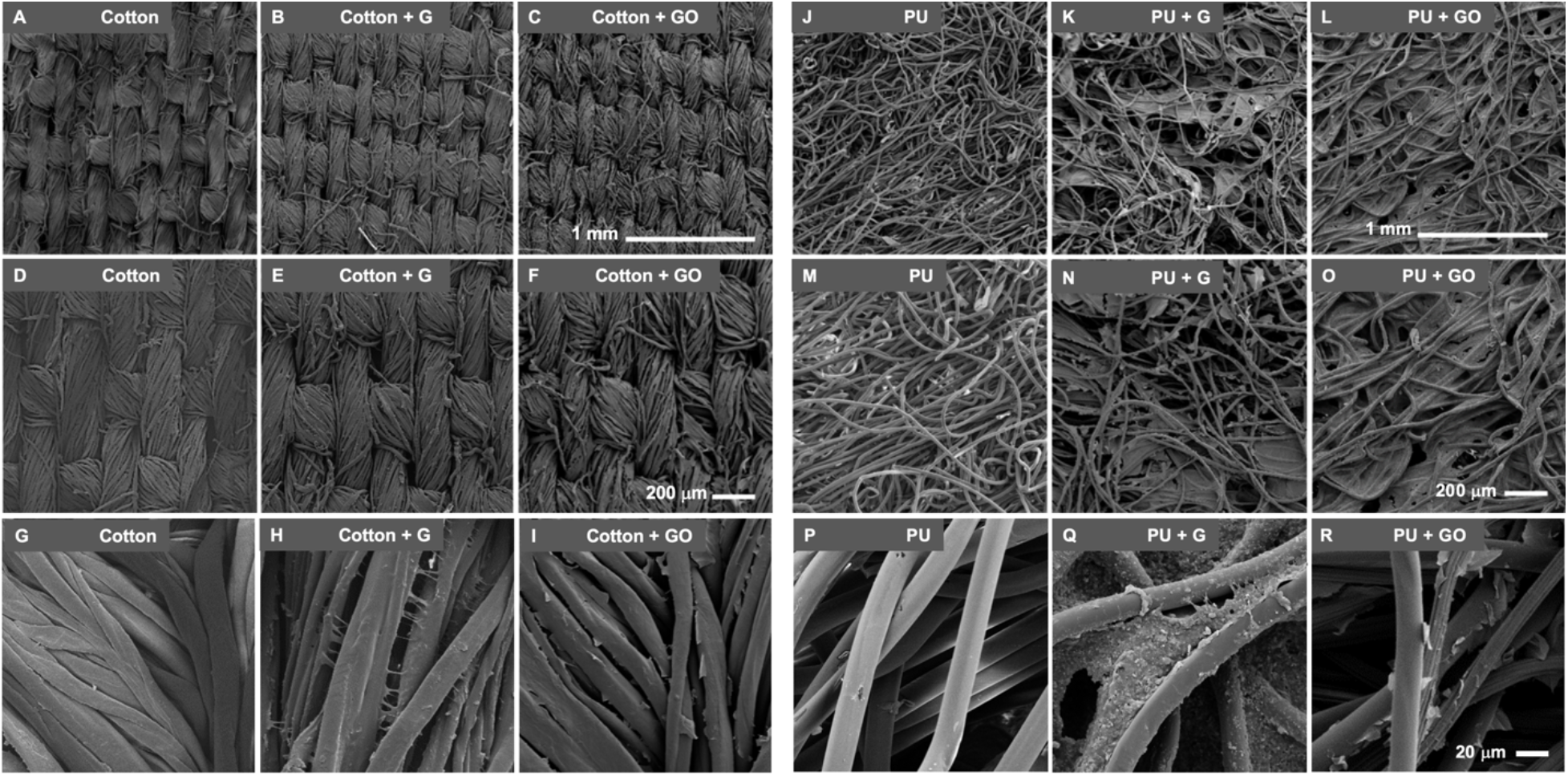
Scanning electron microscopic (SEM) images of Graphene (G) and Graphene oxide (GO) functionalized materials. Representative SEM images of cotton, cotton + G, and cotton + GO at 60X (A, B, and C, respectively), 100 X (D, E, and F, respectively) and 750X (G, H and I, respectively) are shown. Similar images are shown for PU, PU + G, and PU + GO at 60X (J, K, and L, respectively), 100X (M, N, and O, respectively) and 750X (P, Q, and R, respectively).

Two experimental paradigms were used to test protection against SARS-CoV-2 infection: flow filtration and static incubation. The flow and static experimental steps are shown in Figure 3. For the flow study, we generated a custom 3D-printed sample holder for each type of material (Fig. 3A). The sample holder was designed to allow viral suspension filtration through materials used as filter membranes. SARS-CoV-2 suspensions were filtered through each material with or without G/GO functionalization and eluates were collected then used to infect VERO cells per the scheme shown in Fig. 3B.

**Figure 3.**
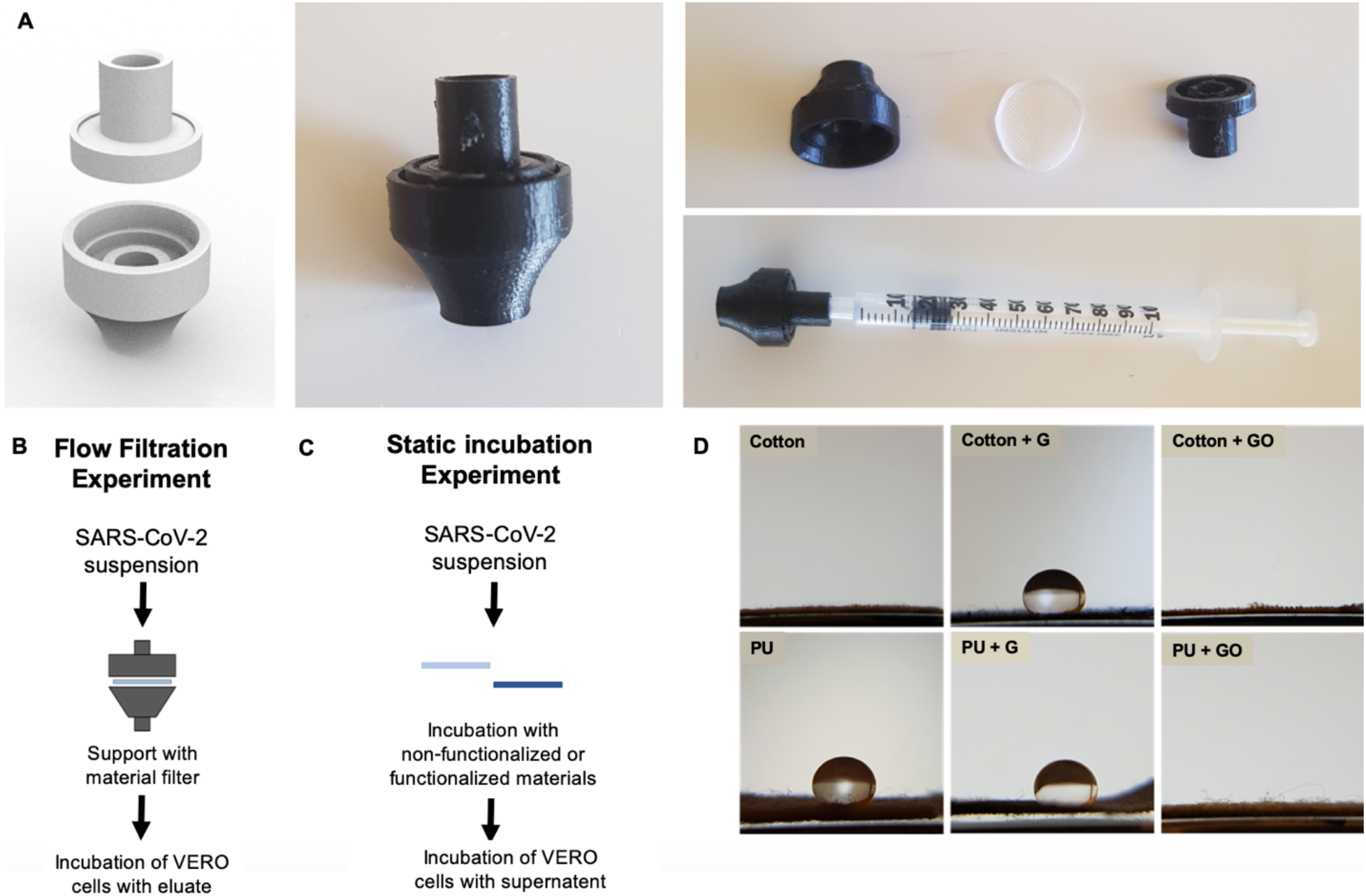
Schematic representation of the experimental design and materials used to assay antiviral properties of non-functionalized and Graphene (G) or Graphene oxide (GO) functionalized materials. (A) A custom 3D-printed device was used to hold the non-functionalized and G- or GO-functionalized materials to filter SARS-CoV-2 viral particle suspensions. (B) In the flow filtration experiment, the viral suspension was filtered through material held in the custom support and the post-filtration eluate was used to infect VERO cells. (C) In the static incubation experiment, viral suspensions were directly incubated with the test materials and the supernatant used to infect VERO cells. (D) Images show the hydrophobicity of each material before after functionalization with G or GO.

In the static testing paradigm, viral particles suspended in medium were incubated G or GO functionalized materials then the medium was used to incubate VERO cells (Fig. 3C).

Remarkably, integration of G or GO into either of the test materials resulted in a highly significant decrease in viral cytotoxicity under both experimental conditions, consistent with the findings with soluble free GO above. As shown in Fig. 4A and B, when eluates flowed through the materials were compared, VERO cell viability was increased by either G or GO integration for both materials suggesting reduction of viral load in the eluate, likely through trapping by the G/GO-integrated material. Very similar results were found for the static experiment (Fig. 4C and D) wherein G or GO functionalized materials were incubated in cell culture medium before it was placed on VERO cells.

**Figure 4.**
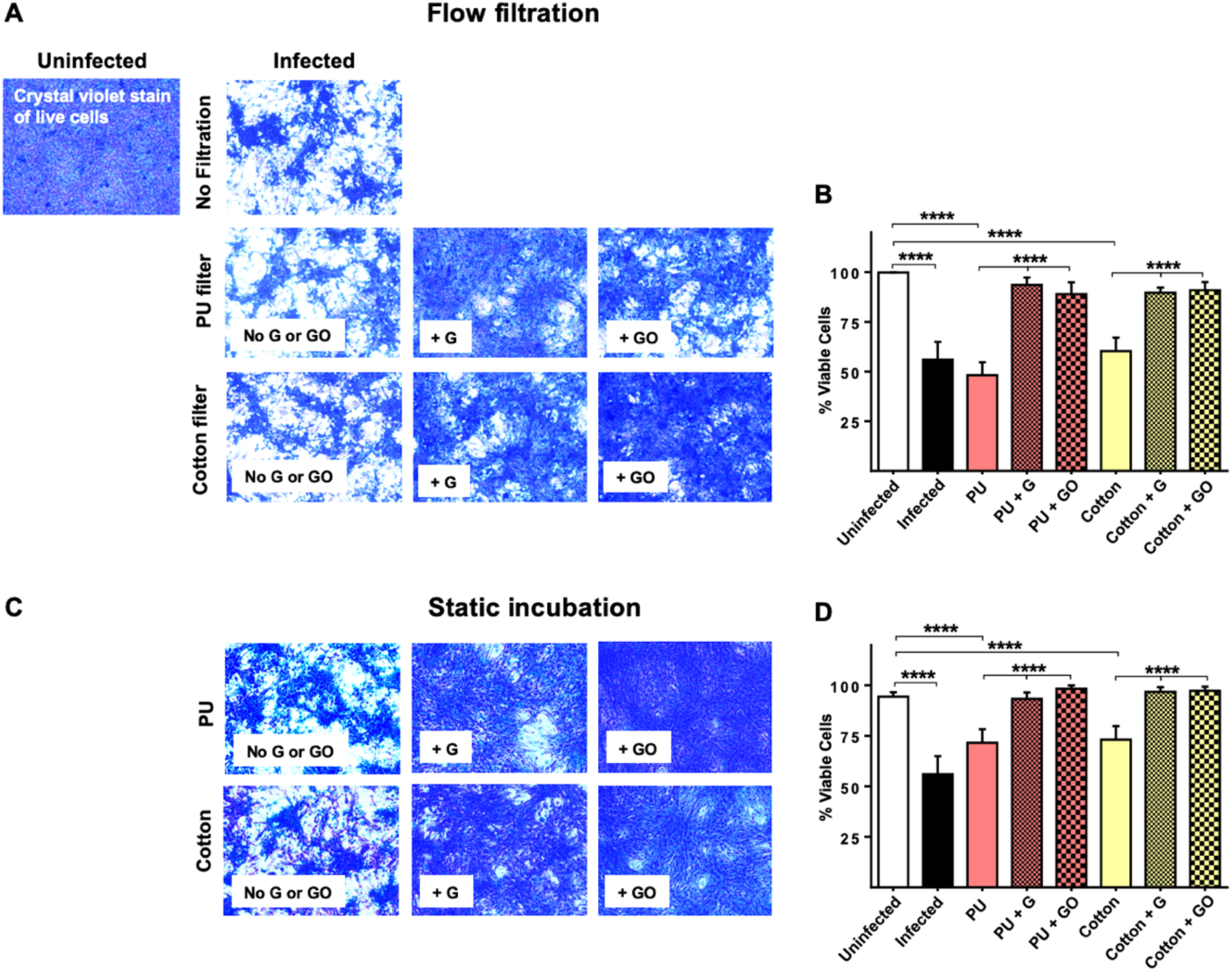
Graphene (G) and Graphene oxide (GO) functionalized materials inhibit viral infection of VERO cells. In the flow filtration experiment, 1 mL of ~10^5^ SARS-CoV-2 viral particles/mL was injected into a custom printed device holding the test materials, flowed through the materials and post-filtration eluates collected. Equal volumes of post-filtration eluates were used to test the ability of functionalized materials to prevent viral infection of VERO cells by (B) Crystal violet staining for cell viability and (C) quantification using ImageJ software. In the static experiment, 0.1 mL of a suspension containing ~10^5^ particles/mL of SARS-CoV-2 was incubated with each material (1×1 cm^2^). After a 2 hour incubation, materials were transferred to tubes containing 5 mL of fresh cell medium and vortexed for 5 seconds × 5; 0.1 mL of each supernatant was used to infect VERO cells. (D, E) Cell viability was detected by using Crystal violet staining. Data graphed as the mean with SD. All experiments were analyzed by using one-way ANOVA comparison tests followed by Tukey’s correction (p < 0.05 = *; p < 0.01 = **; p < 0.001 = ***).

To confirm the functionalized materials themselves are biocompatible and not cytotoxic, A549 pulmonary tumor cells were incubated with cell culture medium exposed to graphene functionalized materials (Fig. 5A and B) and showed no decrease in viability.

**Figure 5.**
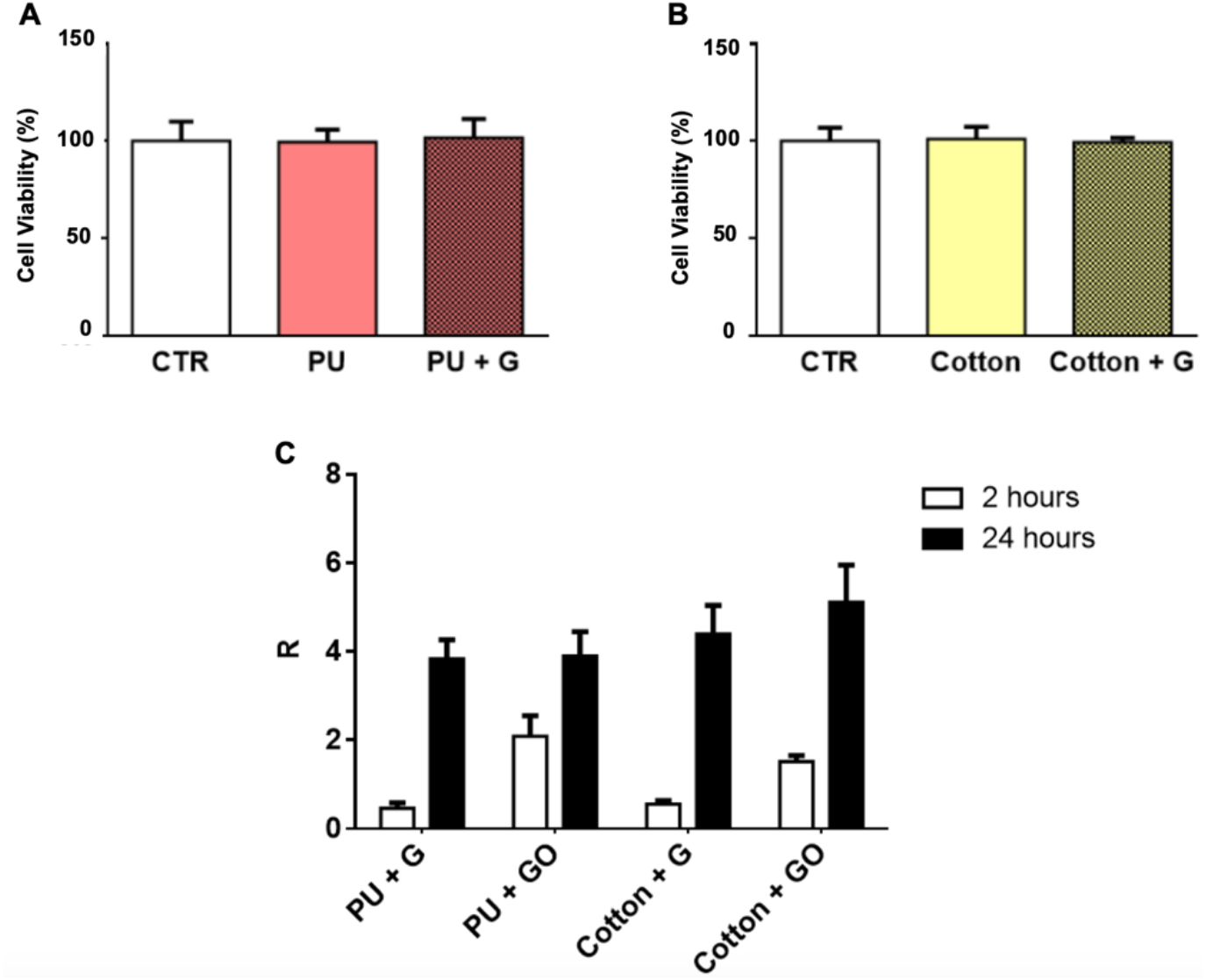
Biocompatability of functionalized materials and anti-bacterial effects. The viability of A549 cells as measured with MTT assay after incubation with functionalized (A) PU-exposed and (B) cotton-exposed supernatants are shown. Quantifications are averages of at least 3 replicates. The viability of cells after incubation with medium exposed to G or GO-loaded materials is not affected. (C) Antibacterial efficacy is shown, determined by calculating the R value for *E.coli* for each sample. Data graphed as the mean with SD.

Material functionalization can also protect against transmission of bacteria. A recent study demonstrated that during surgical operations, the bacterial count on surgical masks originating from the surgeon’s skin increases significantly after 2 hours of usage (*36*). Therefore, we also tested the protective effects of G-integrated materials on *E. coli* growth. After 2 hours, *E. coli* growth was reduced on PU-G and cotton-G with a R value of ~ 0.5 for both; R values were higher for GO-compared to G-functionalized materials (Fig. 5C). The R value is essentially the difference between viable bacteria in an untreated culture and a treated culture, thus if fewer bacteria are viable after a given treatment, the R value is increased. After 24 hours, the R value was 3.8 and 4.4 for PU + G and cotton + G respectively, and 3.8 and ~5 for PU + GO and cotton + GO, respectively; indicating a significant inhibition of bacterial growth by both functionalized materials (Fig. 5C).

While the promising results presented here point to the benefits of either G or GO in PPE material, G and GO are not interchangeable. The hydrophilicity of the material is significantly increased with GO, compared to the hydrophobic G (Fig. 3D). Given the hydrophobicity requirement of face masks, G might be considered the first choice for PPE material functionalization. On the other hand, the effects of GO might be useful in applications in solutions such as molecular diagnostic devices or water treatment, as discussed above.

The findings presented here support the further development of integration of graphene, specifically hydrophobic G, into face mask materials. Face mask use, now encouraged throughout the world where COVID-19 is present, has the potential to reduce viral transmissibility, preserve healthcare capacity, and prevent a second wave of infections (*37*). Graphene and GO nanomaterials present a critical opportunity to increase face mask efficacy and bring the world closer to the goal of stopping the spread of SARS-CoV-2.

## Data Availability

All data is included in the manuscript.

## Acknowledgments

We thank Patricia Spilman of ImmunityBio, LLC for editing this manuscript.

## Funding

Work presented in this paper has been partially supported by Directa Plus Srl and Università Cattolica del SC.

## Author contributions

This study was designed by FDM, VP, GD, MSali PS-S and MP. Infection experiments and related assays were performed by FDM, IP and AS. Bacteria-based experiments were performed by VP and AA. Electron microscopy was performed by VP and AA. Cell viability experiments were performed by FD and GP. Data analysis and figure preparation was conducted by FDM, VP and GB. FDM, VP, GD, MSali, PS-S and MP wrote the paper. MDS and MSanguinetti discussed all the results presented and revised the paper. LGR and GC developed and produced graphene nanoplatelets and realized G material functionalization. MP and VP realized GO material functionalization. All authors read, critically revised and approved the final manuscript.

## Materials and Methods

Graphene nanoplatelet (G) and Graphene oxide (GO) sources

G (G+, Directa Plus) and GO (GO, GrapheneA) were used for all experiments. Commercial materials were used to ensure consistency between experiments. G+ is produced according to a proprietary patented technology that involves three different steps: expansion, exfoliation, and drying (*38*). GO water dispersion (4 mg/mL) synthesis was performed using the Hummers’ method (*39*). In the following table, the main parameters for each sample are summarized. Full characterization of these nanomaterials is reported elsewhere (*40, 41*).

|  | G | GO |
| --- | --- | --- |
| ID/IG (Raman spectroscopy) | 0.127 | 1.1 |
| Thickness (Atomic Force Microscopy) | 4-6 layers | 1 layer |
| Lateral Dimension (Atomic Force Microscopy) | 3μm | 1μm |
| C/O atomic ratio (X ray Photoelectron Spectroscopy) | -- | 1.9 |

### Determination of Graphene oxide (GO) activity in solution against SARS-CoV-2 virus

To assay the ability of GO to trap SARS-CoV-2, we incubated previously titered SARS-CoV-2 particles in suspension with GO diluted in phosphate buffered saline (PBS) supplemented with 1% penicillin – streptomycin. Briefly, 0.06, 0.12, 0.25, and 0.5 mg/mL of GO were incubated with SARS-CoV-2 virus (~10^5^ virus particles/mL). Incubation was performed at 37°C with agitation (100 rpm). Two hours later, GO was removed by centrifugation (14,000 rpm, 5 minutes) and supernatant was used to infect VERO cells (Figure 1A). The cytopathic effect of SARS-CoV-2 was evaluated 72 hours after the infection including full-spectrum light microscopy and image collection for quantification of area covered by viable cells labelled with crystal violet staining. Further infection evaluation was done by labeling of SARS-CoV-2 particles with an anti-spike antibody and performing lactate dehydrogenase (LDH) assay, all described below.

### VERO cell culture

African green monkey kidney (VERO) epithelial cells (ATCC CCL-81) were cultured in Dulbecco’s Modified Eagle’s Medium (DMEM) supplemented with 10% inactivated fetal calf serum (FCS) (EuroClone, Milan, Italy), 1 mM glutamine (EuroClone, Milan, Italy), 1% streptomycin – penicillin antibiotics (EuroClone, Milan, Italy) and incubated in a humidified atmosphere (5% CO_2_ at 37°C) as reported elsewhere (*42*). Cells were washed with sterile warm phosphate buffered saline (PBS), trypsinized and counted. Cells were replated in 48-well plates (Wuxi NEST Biotechnology Co., Ltd, China) at 7 × 10^4^ cells/mL. Cells were infected with SARS-CoV-2 virus when > 90% confluent monolayer was observed comprising approximately 1 × 10^6^ cells/well after 72 hours.

### VERO cell infection

To initiate infection, cells were washed with sterile warm PBS and then infected with 0.1 mL of solution containing SARS-CoV-2 (~10^5^ virus particles/mL). Cells were incubated for two hours at standard atmosphere conditions (37 °C, 5% CO_2_), then the infection solution was removed and replaced with fresh DMEM medium supplemented with 2% FCS, 1 mM glutamine, and 1% streptomycin – penicillin as above. Cells were incubated and infection status was evaluated daily. All experiments that involved SARS-CoV-2 manipulation was carried out in Biosafety level 3 laboratory (BSL3) in the Institute of Microbiology of IRCCS – Fondazione Policlinico Gemelli.

### Immunofluorescent (IF) labelling of SARS-CoV-2 viral particles

IF was performed to assess viral replication in VERO cells. Cells were fixed by using 4% paraformaldehyde for 30 minutes. After three washes, fixed cells were permeabilized (0.02% Triton X-100 in PBS) and a blocking step was performed by using PBS supplemented with 0.3% bovine serum albumin (BSA) (*43*). SARS-CoV-2 viral particles were labelled with monoclonal rabbit anti-Spike S1 subunit antibodies (Novusbio, clone CR3022), the plate was incubated for 3 hours at room temperature, and after washes with PBST, incubated with secondary anti-rabbit IgG – FITC labelled antibodies (Ref: 65-6111/ lot: UG285467, Invitrogen). The fluorescent signal was detected by using a Nikon eclipse TS100 and images used for quantification of the signal as described below.

### Cell viability determination by lactate dehydrogenase (LDH) assay

To evaluate cell viability, LDH levels in cell culture media supernatents were determined (*30–32*). Each supernatant was diluted according to the LDH kit manufacturer’s instructions (LONZA) before incubation with the substrate. Thirty minutes later absorbance at 450 nm was measured with a plate reader (BioRad).

### Crystal violet staining

Crystal violet labels the DNA of live, adherent cells and was used to quantify viable cells. Cells were fixed as described above and then stained by using Crystal violet for 30 minutes. After incubation five washes were carried out and images of random fields for each condition were acquired and the stain signal quantified as described below.

### Image analysis

Images were analyzed using open-access ImageJ software version 1.47v (NIH, USA). Every set of .tiff images corresponding to crystal violet staining were analyzed through the “*Process>Batch>Macro tool”*. Each image was converted to 8-bit image. Minimum and Maximum thresholds were manually set for each batch of images to correctly convert areas to white and black, respectively. Prior to perform the “*Measure*” tool of ImageJ, images were processed with the *"Smooth"* and *"Convert to Mask*". The fraction of the area covered by cells is then automatically stored in the results file. IF images were generated by merging image of cells acquired with direct light and the corresponding image acquired with 476 nm light (UV) by ImageJ software built-in tool *“Image>Color>Merge”* using the grey and green channels, respectively.

G and GO material functionalization and analysis of viral neutralization effects

### Functionalization of materials

Polyurethane (PU) and cotton materials were functionalized with G+ or GO as previously described (*44, 45*).

### Scanning Electron Microscopy (SEM)

SEM was performed to evaluate graphene and GO distribution on materials. A piece of each material was cut (1×1 cm) and sputter-coated with platinum then imaged with SEM Supra 25 (Zeiss, Germany). Images were acquired at several magnifications.

### Antiviral effects of materials functionalized with G or GO under flow filtration

A 3D printed custom sample holder was produced using Ultimaker S3 to hold material samples. SARS-CoV-2 infection solution was prepared as indicated above and 1 mL was filtered through the device containing the material. Positive and negative controls are represented by infection solution and sterile PBS passed through empty filters, respectively. 0.1 mL of each eluate was used to infect VERO cells as indicated above. The cytopathic effect was evaluated at 72 hours after the infection.

### Antiviral effects of materials functionalized with G or GO in static conditions

The antiviral property of materials was evaluated following ISO18184 procedures. 0.05 mL of PBS containing SARS-CoV-2 virus was put on the surface of a 1×1 cm^2^ textile TNT and cotton and corresponding graphene functionalized materials. After two hours of incubation, each material was recovered in a new tube containing 5 ml of DMEM supplemented with 2% inactivated FCS, 1 mM glutamine, 1% streptomycin – penicillin antibiotics. Additionally, 0.1 mL was used to wash and to collect infection solution in the wells containing the materials. Each tube containing infected material was vigorously vortexed five times and 0.1 mL was used to infect VERO cells as described above. The cytopathic effect was monitored daily by visual inspection.

### Biocompatibility of functionalized materials

To assess if the cotton or PU materials functionalized with GNP or GO were toxic to cells in the absence of virus, A549 lung cancer cells (ATCC) were incubated with material-exposed medium. A549 cells were maintained in DMEM (Sigma-Aldrich) supplemented with 10% fetal bovine serum (FBS, EuroClone), 2% penicillin-streptomycin (Sigma-Aldrich) and 2% L-glutamine (Sigma-Aldrich). Cells were cultivated in T75 flasks and kept at 37°C in 5% CO_2_ humidity. Material biocompatibility was evaluated according to ISO 10993. Material pieces (2 × 2 cm) were incubated in 10 mL of complete medium at 37°C for 24 hours. Meanwhile, cells were seeded on 96-well (Corning) at a density of 10^5^cells/mL, and kept at 37°C in 5% CO_2_ humidity for 24 hours. After incubation, supernatant in the multiwell was aspirated and replaced with 100 μL of culture medium incubated with functionalized materials. Cells were then kept at 37°C in 5% CO_2_ humidity. After 24 hours, MTT assay (3-(4,5-dimethylthiazol-2-yl)-2,5-diphenyltetrazolium bromide (Invitrogen, Life technologies, Italy)) was performed. Briefly, culture medium was removed and replaced with fresh medium containing 12 mM MTT. Cells were incubated for 4 hours at 37°C in 5% CO_2_ humidity. After incubation, 100 μL of component B (1 mg of sodium dodecyl sulfate in 10 mL of 0.01 M HCl) per 100 μL of medium was added to each well, and incubated for 16 hours. Absorbance was read at 570 nm with a microplate reader (Cytation3, Biotek) and results were normalized by control cells.

### Assessment of functionalized material antibacterial properties

*E. coli* (ATCC 25922) was used to perform tests of antibacterial effects of materials according to ISO 20743:2013 using a colony plate count method. Cotton or PU without G or GO were used as control materials to validate the growth condition of test bacteria and validate the test.

Bacteria were grown in sterile Luria-Broth (LB) medium at 37 °C overnight. A sub-inoculum of the bacteria was grown until a logarithmic phase of growth was achieved and diluted to a concentration of 10^5^ cells/mL. Material test specimens were cut in pieces (2 cm × 2 cm) and incubated with 200 μL of bacterial suspension and incubated at 37°C. Bacteria were retrieved from each specimen at several time points to count colony forming units (CFU) on LB agar plates. Antibacterial efficacy is expressed with value R which is obtained by R=U_T_-A_T_. Ut is the average of the common logarithm of the number of viable bacteria, in cells/cm^2^, recovered from the untreated test specimens after 24 h; A_T_ is the average of the common logarithm of the number of viable bacteria, in cells/cm^2^, recovered from the treated test specimens after 24 h. All tests were performed in triplicate.

### Statistical analysis

All experiments were replicated at least three times. Microsoft Excel (2010) and Prism6 software (GraphPad) were used to compile and analyze data. All data were expressed as mean with SD and analyzed by one-way ANOVA comparison tests followed by Tukey’s correction.

## Notes

### Competing Interest Statement

Authors Giulio Cesareo, Laura Rizzi, and Patrick Soon-Shiong have an interest in or are employees of Directa-Plus, which makes graphene. Their COIs are attached.

### Funding Statement

Directa-Plus Srl
Universita Cattolica del SC

### Author Declarations

This study did not involve human subjects, thus IRB oversight is not applicable

### Summary of Updates

Massimiliano Papi is now listed as a corresponding author; Giulio Cesareo is removed as corresponding author. Minor edits in contribution section (grammar only).

